# Climate-Informed Deep Learning for Spatio-Temporal Forecasting of Climate-Sensitive Diseases

**DOI:** 10.64898/2026.03.20.26348930

**Authors:** Geletaw Sahle Tegenaw, Mizanu Zelalem Degu, Worku Birhanie Gebeyehu, Asaye Birhanu Senay, Janarthanan Krishnamoorthy, Tomas Ward, Gizeaddis Lamesgin Simegn

## Abstract

**Background:** Effective public health planning and intervention strategies necessitate an understanding of the temporal and geographic distribution of disease incidences. This requires robust frameworks for disease incidence forecasting. However, due to variations in cases and temporal dynamics, grasping the distinct patterns of climate-sensitive diseases poses significant challenges, including identifying hotspots, trends, and seasonal variations in disease incidence. Furthermore, although most studies focus on directly predicting future incidence using historical patterns and covariates, a significant gap remains between methodological proliferation marked by diverse architectures, where models are trained and validated on benchmark datasets that are standardized and statistically stable, and epidemiological reality, which is often characterized by irregular, sparse, and highly skewed data, as well as rare but high-magnitude or bimodally distributed incidences. Hence, traditional end-to-end approaches that directly map climate and disease data often fail in these data-scarce settings due to overfitting and poor generalization. To understand disease epidemiology and mitigate the impact of incidence, we analyzed a decade of retrospective datasets in Ethiopia to examine how climate and weather conditions influence the incidence or spread of climate-sensitive diseases, including malaria and dysentery. In this study, we proposed a two-stage hybrid framework, a climate-informed disease prediction model, to forecast the likelihood of disease incidences using decades of climate and weather data. First, deep learning was applied to capture latent weather dynamics. Then, a hurdle model using Extreme Gradient Boosting (XGB) was designed for zero-inflated incidence data, combining XGBClassifier to predict incidence and XGBRegressor to estimate its size, based on weather dynamics to forecast disease incidence. Our proposed multivariate climate-driven disease incidence model incorporates both spatial (elevation, coordinates) and temporal (year, month) factors, along with key weather parameters (precipitation, sunlight, wind, relative humidity, temperature) to predict the likelihood of multiple diseases occurring in each area, serving as a foundation for future disease incidence predictions in the region. Out of 72 evaluated experiments across four categories and six targets, we found that the Transformer model showed highest number of statistically significant wins (n=18, 25.0%) comparison with Long Short-Term Memory (LSTM) (n=9, 12.5%) and the Temporal Convolutional Neural Network (TCN) (n=5, 6.9%) at climate variable forecasting using Pairwise Model Comparison Diebold-Mariano Test. The hurdle model that combines XGBClassifier and XGBRegressor outperformed the baseline in both Malaria and Dysentery forecasting. Error stratification revealed that the hurdle model provided the greatest benefit during incidence periods, as indicated by a substantially lower Mean Average Error (MAE) in both incidence and non-incidence periods than the baseline. Our proposed modular pipeline first forecasts climate variables, then predicts disease incidence, thereby enhancing interpretability and generalization in data-sparse settings. Overall, this approach provides a scalable, climate-aware forecasting tool for public health planning, particularly in regions where these diseases are endemic or where climate change may affect their prevalence, as well as in data-scarce settings.

**Author summary:** Effective public health planning and intervention strategies: (I) necessitate an understanding of the temporal and geographic distribution of disease incidences, (II) poses significant challenges due to variations in cases and temporal dynamics, grasping the distinct patterns of climate-sensitive diseases, including identifying hotspots, trends, and seasonal variations in disease incidence, and (III) requires effective model for predicting future incidence using historical patterns and covariates. However, existing models are trained and validated on standardized, statistically stable benchmark datasets. In contrast, real-world epidemiological data are often irregular, sparse, and highly skewed, with rare but high-magnitude or bimodal incidence distributions. To fill this gap, this manuscript presents a two-stage hybrid framework, a climate-informed disease prediction model, to forecast the likelihood of disease incidences using decades of climate and weather data. First, to capture latent weather dynamics, deep learning was applied. Then, a hurdle model using Extreme Gradient Boosting (XGB) was designed for zero-inflated incidence data, combining XGBClassifier to predict incidence and XGBRegressor to estimate incidence size, based on weather dynamics to forecast disease incidence. Our proposed multivariate climate-driven disease incidence model incorporates both spatial (elevation, coordinates) and temporal (year, month) factors, along with key weather parameters (precipitation, sunlight, wind, relative humidity, temperature) to predict the likelihood of multiple diseases occurring in each area, serving as a foundation for future disease incidence predictions in the region.

## Introduction

Climate change is increasingly recognized as a critical driver of infectious disease dynamics, with rising temperatures, erratic precipitation patterns, and extreme weather events amplifying the risk of incidences in vulnerable regions [1–5]. The interplay between environmental factors and disease transmission is particularly pronounced in low-resource settings, where limited healthcare infrastructure and socioeconomic disparities exacerbate public health vulnerabilities [6,7]. In sub-Saharan Africa, for instance, climate-sensitive diseases such as malaria, cholera, and meningitis remain endemic, with their incidence often correlated to seasonal climatic shifts [8–10]. For example, malaria incidence in East Africa has been linked to temperature fluctuations and rainfall variability, with warming highlands enabling vector expansion into previously unaffected regions [11–14] as mosquito vector survival and parasite development rates are temperature-dependent [15]. Similarly, cholera incidences have been linked to sea surface temperature changes and precipitation patterns, seasonal floods, which impact bacterial proliferation and human exposure pathways [16–19].

Effective disease forecasting is crucial for proactive public health interventions, early warning systems, and resource allocation. This requires understanding disease distribution across time and location to enable timely interventions. However, climate-sensitive diseases follow complex patterns influenced by environmental factors, making it difficult to identify hotspots, trends, and seasonal variations. Fluctuations in cases, shifting climate conditions, and evolving transmission dynamics further complicate surveillance and forecasting. Integrating climate data with epidemiological models can potentially enhance prediction accuracy, enabling proactive disease control. Early climate change forecasting is crucial for anticipating vector-borne and waterborne disease risks, allowing for timely interventions and efficient resource allocation.

Traditional disease forecasting models rely on epidemiological surveillance data, demographic information and are implemented using static statistical models such as generalized linear models (GLMs), logistic regression, and autoregressive integrated moving average (ARIMA) models [20–23]. While these models provide valuable foundational insights, they often fail to fully capture the dynamic relationships between climate variability and disease incidence.

With the rise of artificial intelligence, machine learning (ML) and deep learning (DL) approaches have been increasingly adopted for disease prediction. For example, random forest and support vector machines (SVM) have been used for dengue and malaria forecasting, leveraging climate and entomological data to enhance prediction accuracy [24–27]. More recently, recurrent neural networks (RNNs) and long short-term memory (LSTM) models have been employed for time-series forecasting of infectious diseases [28,29], offering improved handling of sequential data and capturing non-linear trends. Despite these advancements, challenges remain in designing robust models that effectively integrate multivariate climate data with multiple epidemiological information. Many existing ML-based approaches are often limited by their focus on a single climate-sensitive disease, as well as by data sparsity, regional variability, and generalization issues that hinder their real-world applicability.

The emergence of transformer-based architectures represents a paradigm shift in time series forecasting [30–34]. These models excel at capturing long-range dependencies and multi-scale patterns, making them particularly suited to climate-disease systems. In climate-sensitive disease systems, where non-linear relationships between environmental drivers (e.g., temperature, humidity) and pathogen prevalence are common, transformers’ ability to process high-dimensional spatiotemporal data offers a distinct advantage. In the realm of climate-related health forecasting, transformer-based models have shown considerable promise. A study employing a Fourier mixed window attention transformer [33] has been introduced to predict dengue fever cases by analyzing local climate data and global climate indicators. This approach helps to capture intricate relationships between climatic factors and disease incidence, facilitating long-range forecasts of up to 60 weeks. Similarly, a method integrating climate variables from multi-sensor remote sensing imagery with advanced machine learning techniques [34] has been proposed for the grapevine disease prediction. This technique demonstrates the potential of transformer-based models in processing complex environmental data to forecast disease incidences in viticulture.

Despite these advances, most existing models adopt end-to-end architectures that directly map climate and epidemiological inputs to future disease incidence. In low and middle-income countries (LMIC), epidemiological surveillance systems are often incomplete and prone to reporting delays, making it impractical to rely solely on end-to-end deep learning models trained directly on disease case counts, as these models tend to exhibit poor generalization, overfitting, and limited trustworthiness among policymakers. On the other hand, climate and environmental variables such as precipitation, temperature, and humidity should routinely be collected at high spatial and temporal resolution through satellite and meteorological networks. However, this is rarely the case in low-resource settings. This limitation constrains the transformative potential of end-to-end deep learning frameworks, which often rely on probabilistic modeling to quantify uncertainty in multi-output forecasts, a critical feature for public health prioritization in data-sparse environments.

Furthermore, sub-Saharan African countries, such as Ethiopia, exemplify the intersection of climate vulnerability and infectious disease burden, characterized by climate data from the meteorological agencies but limited and often incomplete disease incidence records. The varied elevations across Ethiopia create distinct ecological zones, each with unique climate conditions that affect the distribution and transmission of infectious diseases. For instance, malaria, traditionally confined to lower altitudes, has been reported in highland areas due to rising temperatures and the expansion of the habitat range of malaria vectors. Additionally, diseases like dysentery have shown spatiotemporal variations correlating with these microclimates [35]. The convergence of environmental changes and population movements has created conducive conditions for the spread of infectious diseases [36–38]. Developing robust climate forecasting and multi-disease prediction models is essential for addressing the complex interplay between climate variability, ecological factors, and human health. By integrating climate data with epidemiological trends, these models can increase disease surveillance systems, enabling early detection and targeted interventions. Additionally, anticipatory actions, such as early warning systems and community-based interventions, can help mitigate the spread of infectious diseases and strengthen public health resilience in the face of environmental changes.

In general, despite the progress of machine learning and deep learning in disease incidence forecasting, (I) A significant divide remains between methodological proliferation marked by diverse architectures and epidemiological reality. Existing models were trained and validated on benchmark datasets that are abundant, standardized, and statistically stable, yet rarely reflect the irregular, sparse, and biased nature of real-world disease surveillance data; (II) Existing studies focus on directly predicting future incidence using historical patterns and covariates and were successful in learning continuous and not zero-inflated spatiotemporal representations; and (III) Due to variations in cases and temporal dynamics, grasping the distinct patterns of climate-sensitive diseases poses significant challenges, including identifying hotspots, trends, and seasonal variations in disease incidence. In this study, we reformulate the disease incidence problem into a two-stage modeling pipeline. We first fine-tuned a deep learning model to forecast time-series weather variables, which were subsequently used as inputs to disease prediction models. The two-stage hurdle forecasting pipeline was designed for incidence classification (whether an incidence will occur or not) and magnitude regression (the expected magnitude of incidence). A modular climate-informed multi-disease prediction framework utilizing a fine-tuned deep learning-based forecasting model was proposed by separating the task of climate feature model extraction from the tasks of disease incidence modeling to mitigate overfitting in low data settings. By incorporating decadal climate and weather data, including precipitation, temperature, and sunlight exposure, alongside spatial and temporal factors, our approach aims to enhance disease incidence forecasting, enhance interpretability, transferability across regions, and align with policy needs.

## Methods

### Data Sources

We employed a spatiotemporal distribution of climate-sensitive disease incidence dataset in Ethiopia, which contains a longitudinal incidence record of malaria and dysentery diseases between 2010 and 2022/2023 [39]. As mentioned in [39], the main source of data for the study was secondary data collected from the Ethiopian Public Health Institute (EPHI): public health emergency management (PHEM), and the Ministry of Health (MoH), following the approval of the research by the ethical review board of the Jimma Institute of Technology, with the reference number RPD/JIT/172/15.

### Data Preparation and Preprocessing

A set of preprocessing steps was conducted in the historical dataset before modeling the time series deep learning model. Mean imputation was used to fill in the missing values (by filling the most frequent value for non-numeric variables), and Z-score measures to remove outliers. The feature set comprises both feature variables (region, zone, elevation, year, and month) and the target variable based on minimum, maximum, average, and total. Both features and targets are normalized independently using the Min-Max Scaler. The final preprocessed data ready for modelling contains region, zone, station, elevation, location, year, month, perception, relative humidity, maximum temperature, minimum temperature, and wind factors and variables. Furthermore, for each climate variable, we derived four distinct trend datasets: minimum (Min), maximum (Max), average (Avg), and cumulative total (Tot) values from 2010 to 2022 (Figure 1). To represent the distinct physical phenomena (Min, Max, Avg, and Tot), each of these trend categories was treated as an independent modeling target to avoid mixing trends, improve modeling dynamics, and enhance model interpretability. In the end, the dataset was randomly partitioned into training (80%) and test (20%) subsets to ensure independent evaluation

**Figure 1:**
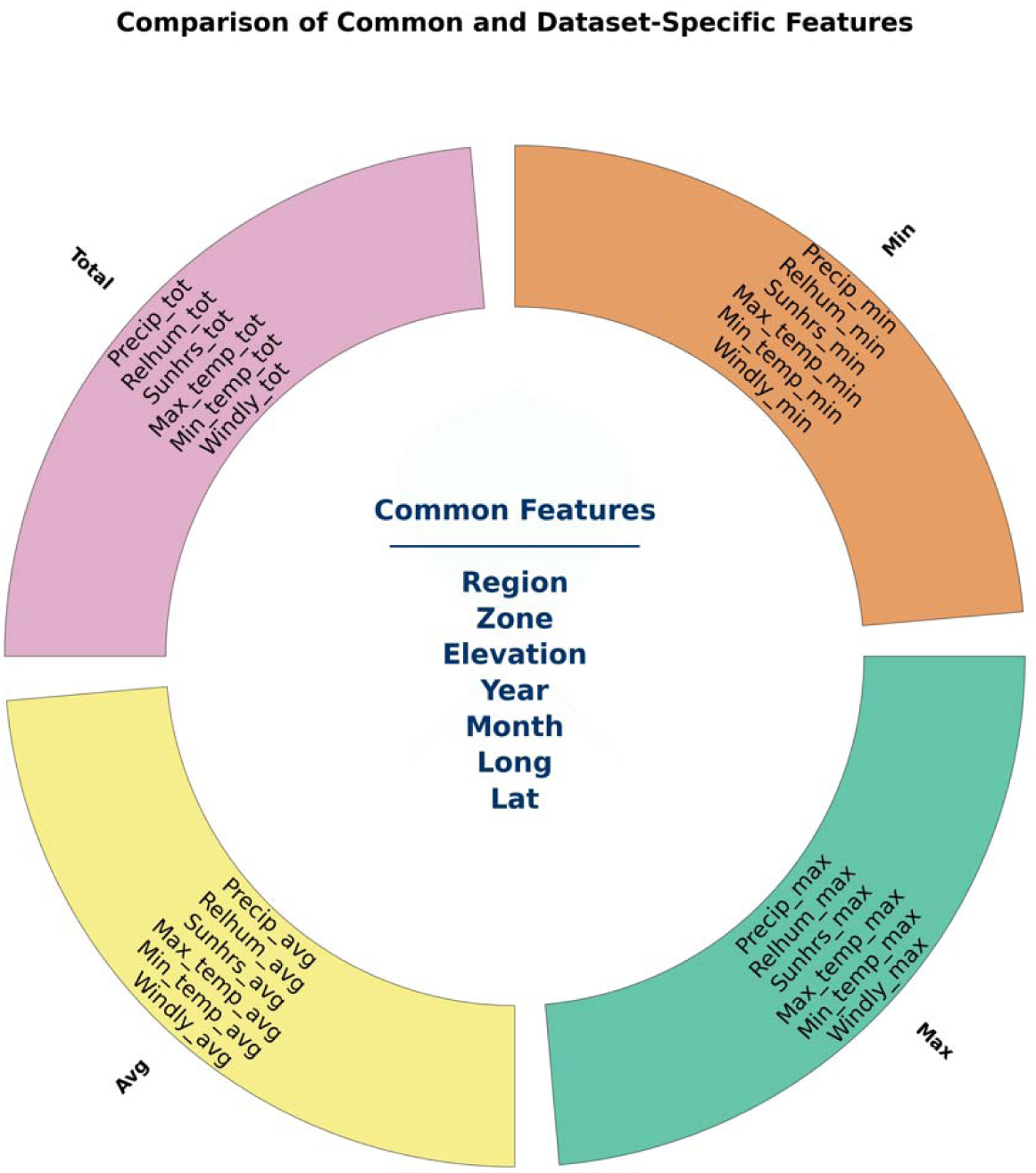
Preprocessed features across each meteorological dataset

### Data Analysis and Modelling

A two-stage modular climate-informed multi-disease prediction framework was proposed for: (I) Forecasting time series climate variables using deep learning, such as perception, relative humidity, maximum temperature, minimum temperature, and wind factors, and (II) Predicting the incidence of Dysentery and Malaria occurrence. The hurdle forecasting pipeline wa designed for incidence classification and magnitude regression. The proposed disease forecasting pipeline is presented in Figure 2, and the framework was designed for utilizing a deep learning-based forecasting model by separating the task of climate feature model extraction from the tasks of disease incidence modeling to mitigate overfitting in low data settings.

**Figure 2:**
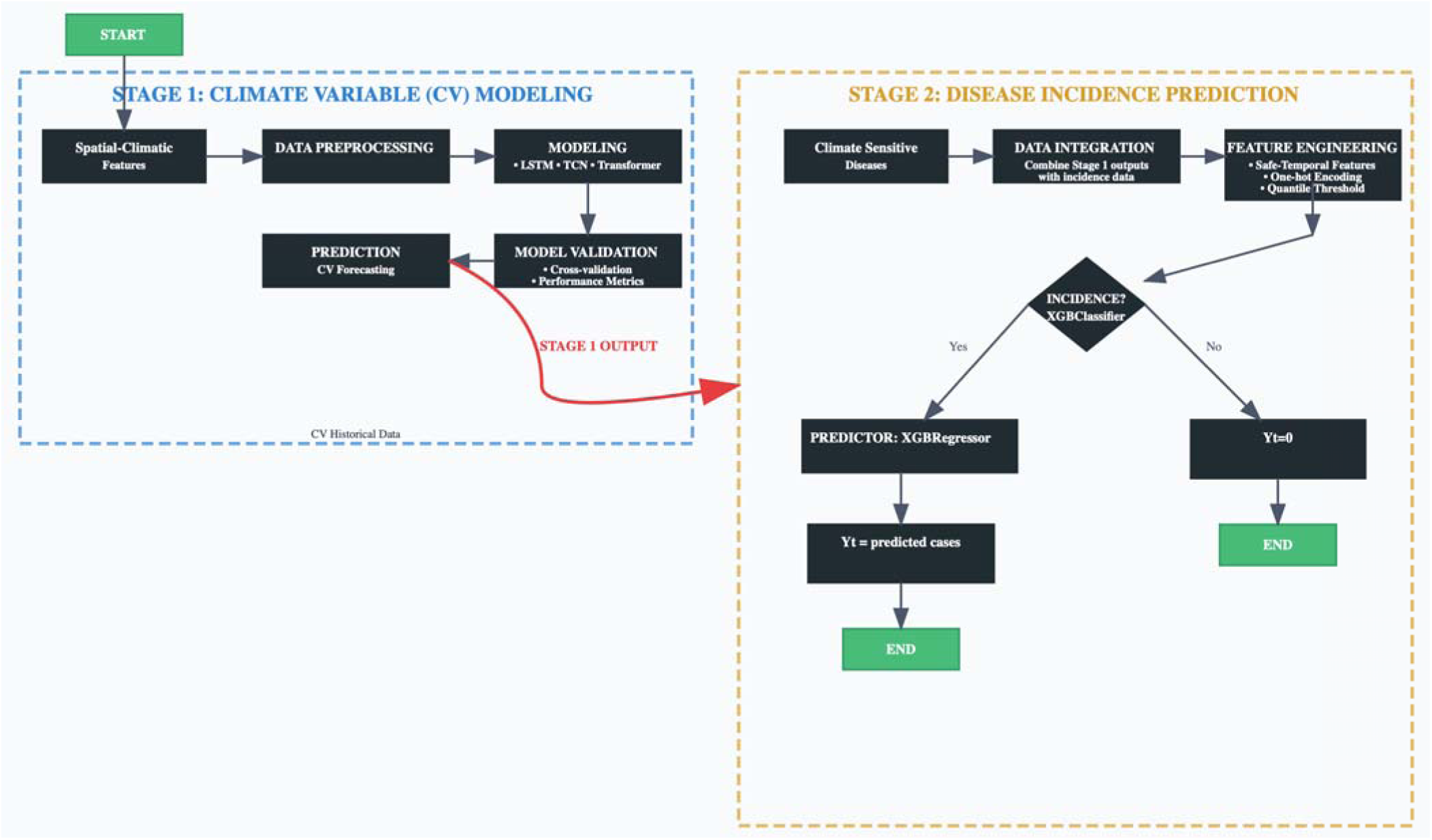
A framework for integrating climate variables into disease forecasting models. Stage 1: Climate Variable Modeling using deep learning. Stage 2: A disease incidence hurdle model (to handle zero-inflated incidence) using latent weather dynamics (i.e., Stage 1 output) and historical disease records, where incidence classification was computed first to classify whether an incidence would occur or not, and then, magnitude regression was computed to estimate the expected magnitude of the incidences.

The analysis and modeling of both climate variable prediction models were conducted based on the normalized features. The normalized features and targets are then horizontally concatenated to create a single data array, which is segmented into sequences using a sliding window of 12 months. Each sequence (with a shape of 12 features) is used to predict the target values at the subsequent time point. The dataset is divided into features and targets.

We employed three forecasting time-series deep learning models via the Darts library [40], including Long Short-Term Memory (LSTM) [41], Transformer [42–43], and Temporal Convolutional Neural Network (TCN) [44]. TCN is a deep learning architecture specifically designed for sequential data like time series, but unlike RNNs (LSTM), it uses convolutional layers instead of recurrent ones. To ensure a fair comparison, all neural models were trained for 100 epochs with a common input chunk length of 24 months. We used fixed seeds, with a random state of 42 for reproducibility. The model utilized historical sequences of 12 months of environmental and contextual features to predict future values for a set of targets. The features include geographic and temporal information (region, zone, elevation, location, year, and month) combined with historical values from the target variables. The final evaluation is performed using standard regression metrics, and results are visualized with scatter plots comparing actual and predicted values.

After training, the model is evaluated on the test set. An evaluation function computes key metrics, including Mean Absolute Error (MAE), Mean Squared Error (MSE), Root Mean Squared Error (RMSE), and the R² score for each target variable. Additionally, scatter plots are generated to visually compare the actual versus predicted values for each target. To provide an overall assessment, we calculated ranks for each model based on its relative performance across four metrics, followed by an average rank score to emphasize the overall effectiveness of the model beyond individual metrics. MAE, MSE, and RMSE measure the size of prediction errors, with lower values indicating better performance. R² indicates the proportion of variance explained by the model and ranges from 0 to 1, with higher values showing a better fit. Finally, to compare forecast accuracy between models, we used the Diebold-Mariano (DM) test, a statistical test used to compare the predictive accuracy of two forecasting models [44, 45], across target variables. Additionally, we visualized error distribution (actual vs predicted) and interpretability.

At the end, following the climate variable prediction model, a disease prediction model of Dysentery and Malaria was trained (Figure 2). The hurdle model was based on Extreme Gradient Boosting (XGB) by combining XGBClassifier and XGBRegressor [46]. The hurdle model was used to predict both the occurrence and size of incidents: (I) a classifier predicts whether a time step is an incident (XGBClassifier), and (II) a regressor estimates the size of incidents only during periods classified as incidents (XGBRegressor). A naive persistence model predicting the previous time step served as the baseline. In this study, incidents were defined as observations above the 20^th^ percentile of the training data within each cross-validation fold. A nested time series split was used, with the outer loop consisting of five splits and 12-month test sets, and the inner loop consisting of three splits for hyperparameter tuning. Grid search explored tree depth, learning rate, and number of estimators. Classifier selection optimized F1-score to balance Precision and Recall for rare incidence; regressor selection optimized R² for magnitude prediction. We evaluated regression performance using R², RMSE, and MAE. Incidence detection performance was evaluated using Precision, Recall, and F1 scores. We also computed incidence-specific MAE and non-incidence MAE to distinguish predictive accuracy between incidence and non-incidence periods.

## Results

### Part I: Climate Variable Modeling

#### Model Performance Across Weather Variable Forecasting Targets

Modeling results of six climate variables based on average, minimum, maximum, and total trends are demonstrated in Table 1. The average trends from 2010 to 2022 shows that: (I) on the precipitation, TCN scored the lowest MAE (0.86), MSE (1.17), and RMSE (1.08), (II) on the relative humidity, LSTM scored the lowest MSE (17.91), and RMSE (4.23), (III) on the sun hours, TCN scored the lowest MAE (0.41), MSE (0.37), and RMSE (0.61), alongside the highest *R²* score (0.61), (IV) on the max temperature, TCN scored the lowest MAE (0.62), MSE (0.58), and RMSE (0.78), alongside the highest *R²* score (0.54), (V) on the minimum temperate, TCN scored the lowest MAE (0.41), MSE (0.24), and RMSE (0.49), and (VI) on the windily, TCN scored the lowest MAE (0.03), MSE (0.002), and RMSE (0.05). Based on the minimum trends, TCN scores the lowest MAE in relative humidity, sun hours, maximum temperature, minimum temperature, and wind, while LSTM scored the lowest MAE in precipitation (Table 1). Whereas, on maximum trends, TCN achieved the lowest MAE, MSE, and RMSE (Table 1). In the case of total trends, the TCN score has the lowest MAE, MSE, and RMSE.

**Table 1.**
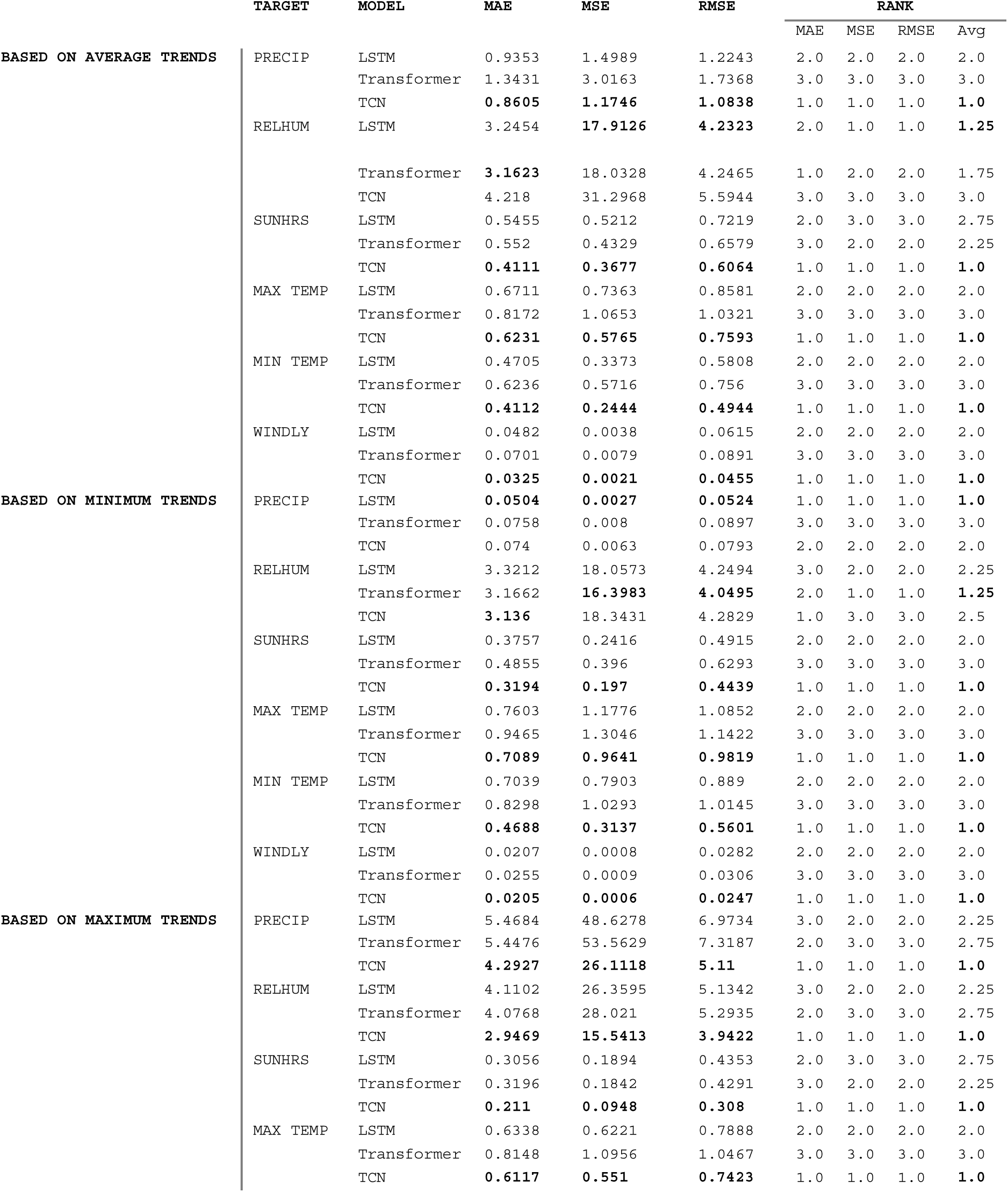
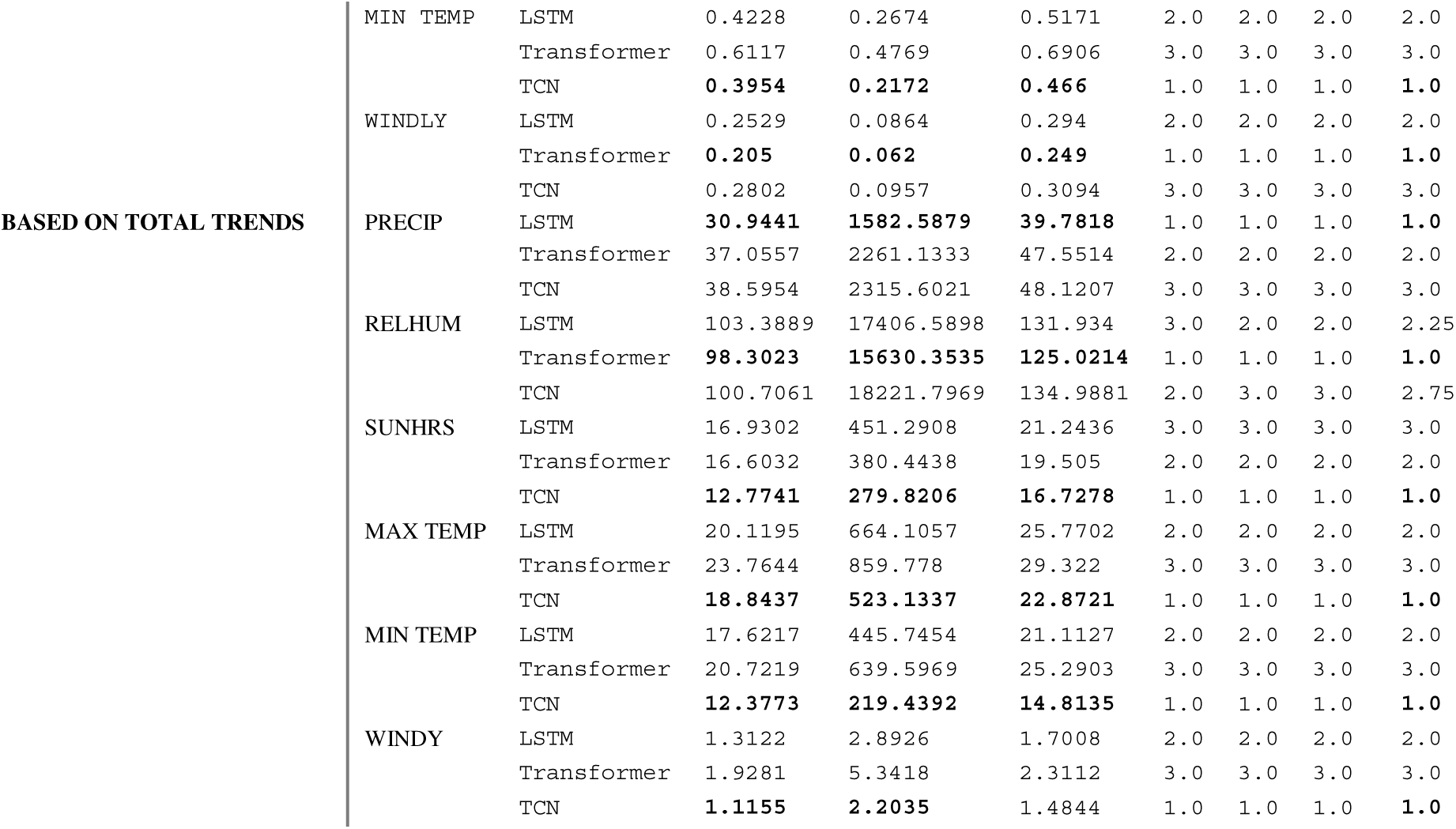
compares the performance of LSTM, Transformer, and TCN models across six climate variables (CV) forecasting targets, including precipitation, relative humidity, sun hours, maximum temperature, minimum temperature, and wind speed-based average, minimum, maximum, and total trends. Metrics include Mean Absolute Error (MAE), Mean Squared Error (MSE), and Root Mean Squared Error (RMSE). The best values for each target and metric are highlighted in bold.

#### Rank-Based Comparative Evaluation

Based on the aggregate assessment ranking of the four metrics (Table 1), we found that TCN leads in predicting climate variables using average trends, including precipitation, sun hours, maximum temperature, minimum temperature, and wind, while LSTM leads for relative humidity. Similarly, on predicting climate variables, TCN leads except in precipitation and relative humidity. Moreover, TCN leads in predicting climate variables based on maximum trends. Likewise, in predicting the climate variable based on total trends, TCN leads except for precipitation.

#### Pairwise Model Comparison using the Diebold-Mariano Test

For average precipitation, the Transformer significantly outperformed the TCN (DM = 2.21, *p* = 0.027). For average relative humidity, TCN significantly outperformed both LSTM (DM =-3.03, *P=0.030*) and Transformer (DM =-3.77, *P<0.001*). While LSTM outperforms Transformer (DM =2.166, *P = 0.030*). Both LSTM (DM =1.999, *P<*) and Transformers (DM =2.17, *P<0.030*) outperform TCN on the prediction of average sun hours. No significant difference was observed in predicting max temperature trends, while in predicting minimum temperature trends, both LSTM (DM =2.211, *P<0.027*) and Transformer (DM =1.961, *P<0.049*) outperform TCN. LSTM significantly outperformed the Transformer (DM =2.399, *P<0.016*) in predicting wind based on average trends. Further information is presented in Table 2.

**Table 2:**
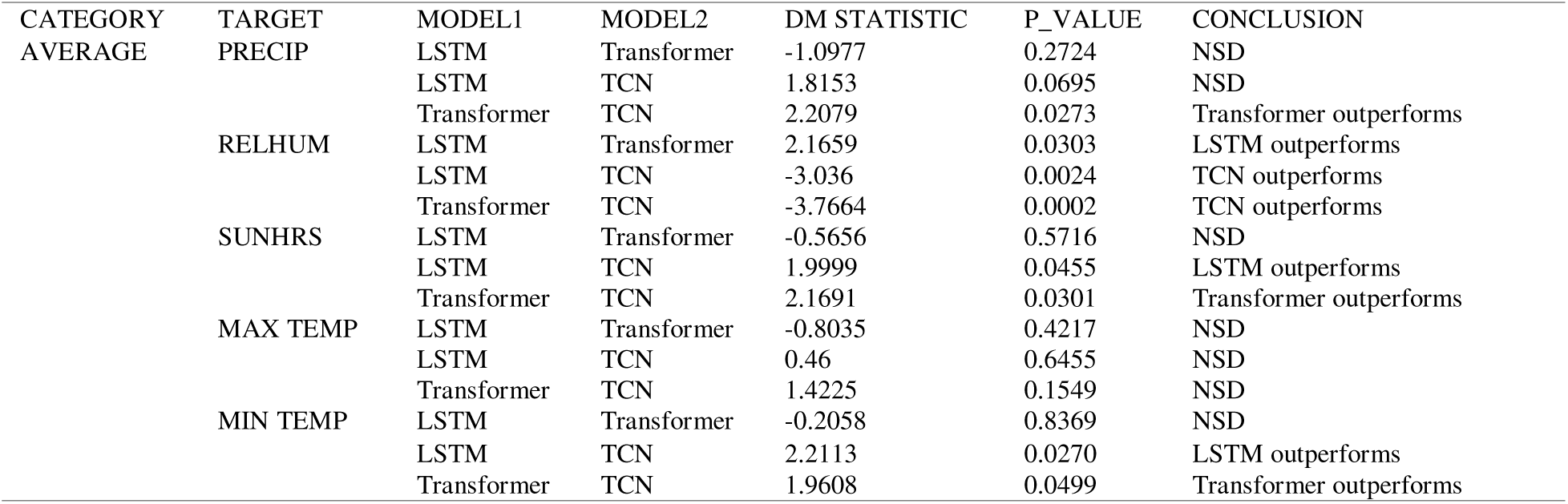
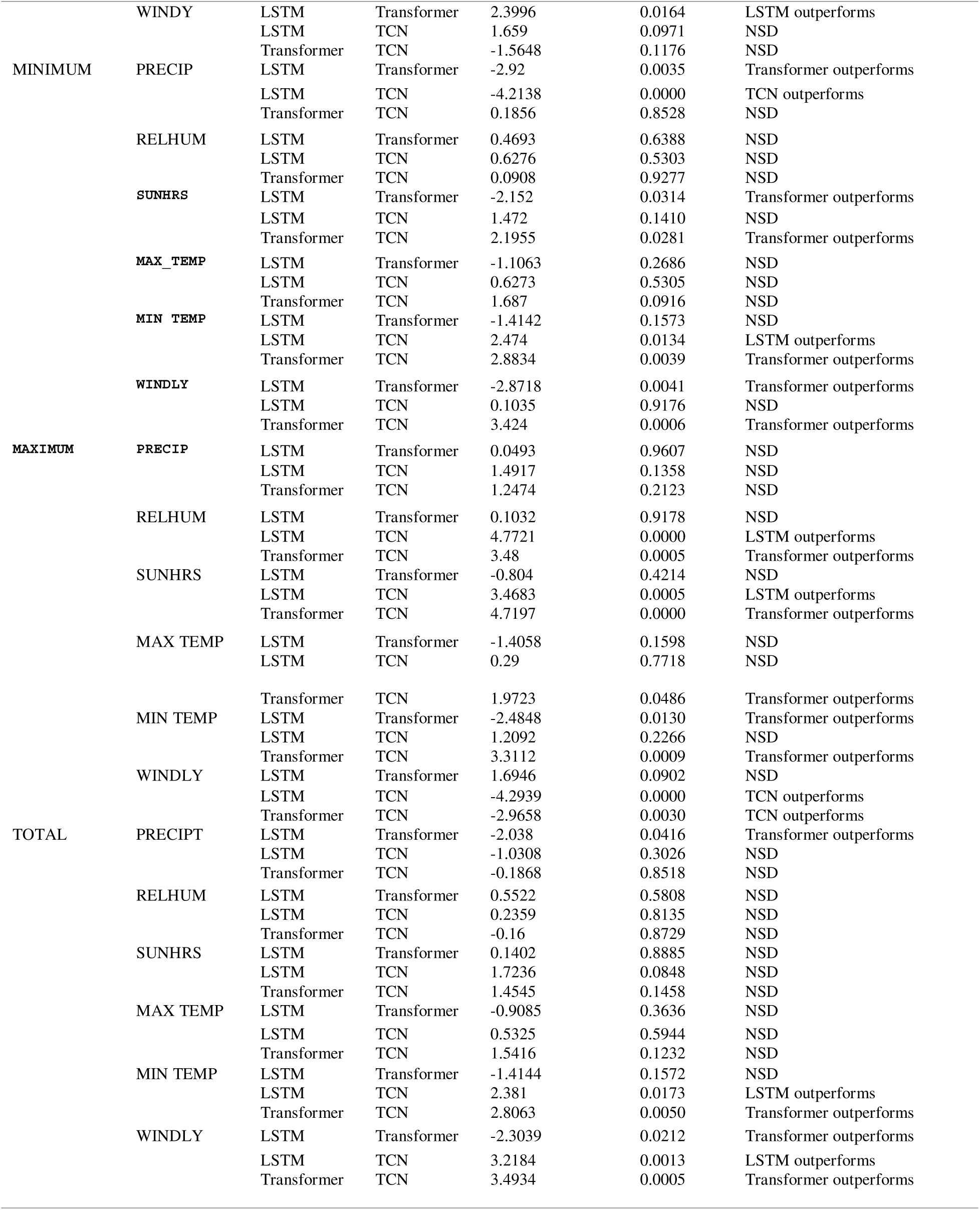
Pairwise Model Comparison via Diebold-Mariano Test. NSD indicates no significant difference.

The transformer significantly outperformed LSTM (DM = −2.920, *P* = 0.004) in predicting precipitation based on the trends of the minimum value. The transformer also significantly outperformed TCN (DM = 2.195, P = 0.0281) in predicting sun hours based on minimum value trends. It also outperformed both LSTM (DM = −2.872, P = 0.004) and TCN (DM = 3.424, P < 0.001) in predicting wind based on minimum value trends.

For predicting precipitation based on maximum value trends, no significant difference was observed (Table 2). Predicting sun hours based on the maximum trends, both LSTM (DM = 3.468, P<0.001) and Transformer (DM = 4.719, P<0.001) significantly outperformed TCN. Transformer outperformed TCN on both maximum temperature (DM = 1.972, P<0.048) and minimum temperatures (DM = 3.311, P<0.001) for predicting based on minimum trends. Whereas TCN outperformed both LSTM (DM = −4.294, P<0.001) and Transformer (DM = - 2.966, P<0.001) in predicting wind based on maximum value trends. No significant difference was observed in predicting precipitation, relative humidity, sun hours, and maximum temperature climate variables based on the total trends, except for minimum temperature and wind. On the wind variable, Transformer outperformed both LSTM (DM = −2.304, P<0.021) and TCN (DM = 3.493, P<0.001), while LSTM outperformed TCN (DM = 3.218, P = 0.001).

#### Actual vs. Predicted Error Distribution Across Trend Categories

Across all four trend categories, average, minimum, maximum, and total, the results consistently demonstrate that the TCN provides the most reliable and accurate predictions. In Figures 3–6, TCN models show tight clustering of predicted values around the ideal y = x reference line, reflected in low MAE and RMSE values across most subpanels. This pattern indicates strong alignment between actual and predicted values and highlights the model’s stability across different types of trend dynamics. In contrast, the LSTM and Transformer models exhibit more variable performance. LSTM performs reasonably well in certain scenarios (e.g., minimum trends) but shows substantial prediction errors in more complex cases, particularly in total-trend estimation, where error values increase significantly. The Transformer model occasionally approaches TCN’s performance in specific panels but overall displays higher error variability and reduced consistency across trend types.

**Figure 3.**
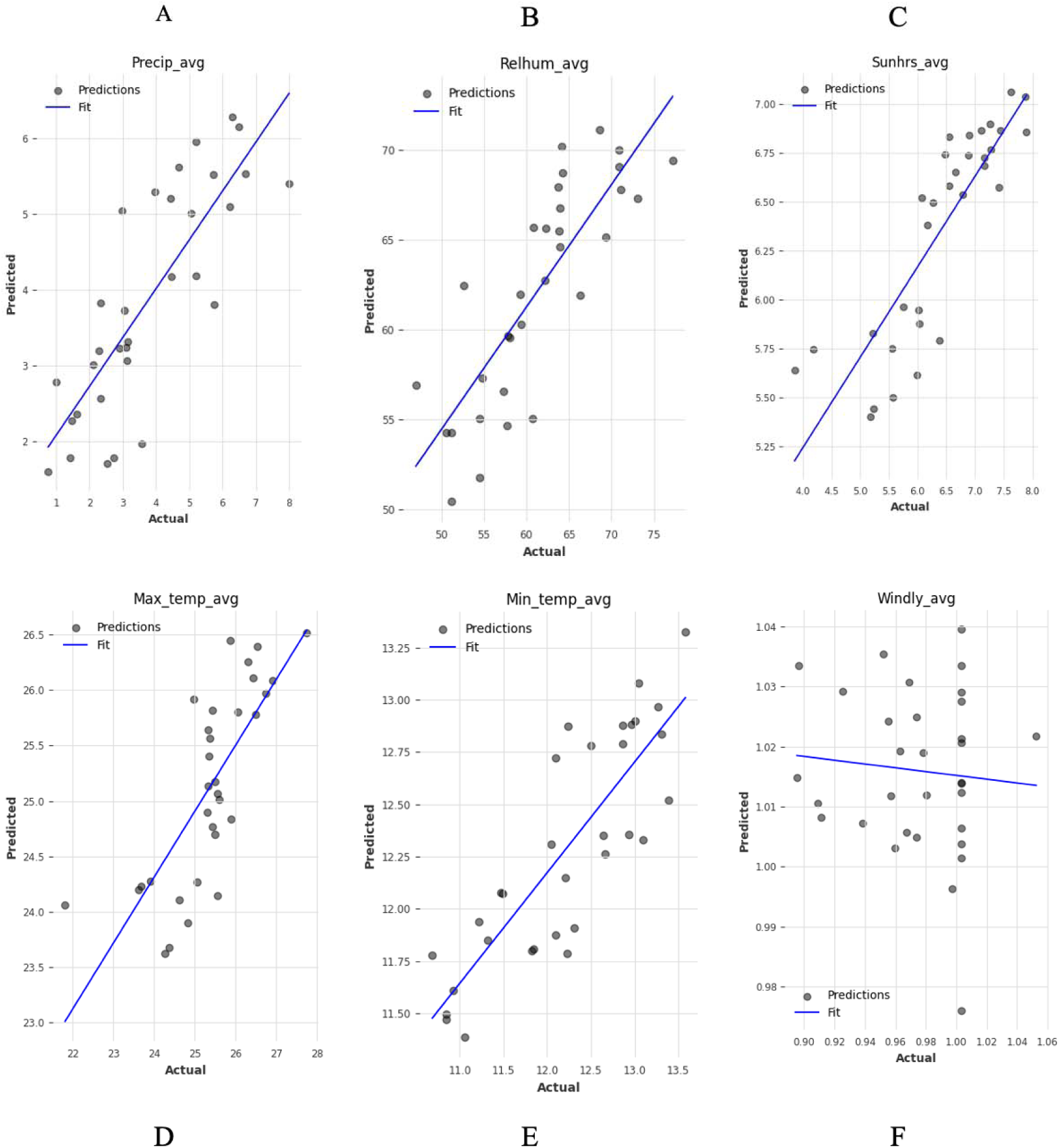
Actual vs. predicted values based on average trends. A) TCN achieved MAE=0.86 & RMSE=1.08. B) LSTM achieved an MAE=3.25 & RMSE=4.23. C) TCN achieved MAE=0.41& RMSE=0.6. D) TCN achieved MAE=0.62 & RMSE=0.76. E) TCN achieved MAE=0.41& RMSE=0.49. F) TCN achieved MAE=0.03 & RMSE=0.04. The black line indicates perfect prediction (y = x). Points clustered near the line indicate accurate predictions.

**Figure 4.**
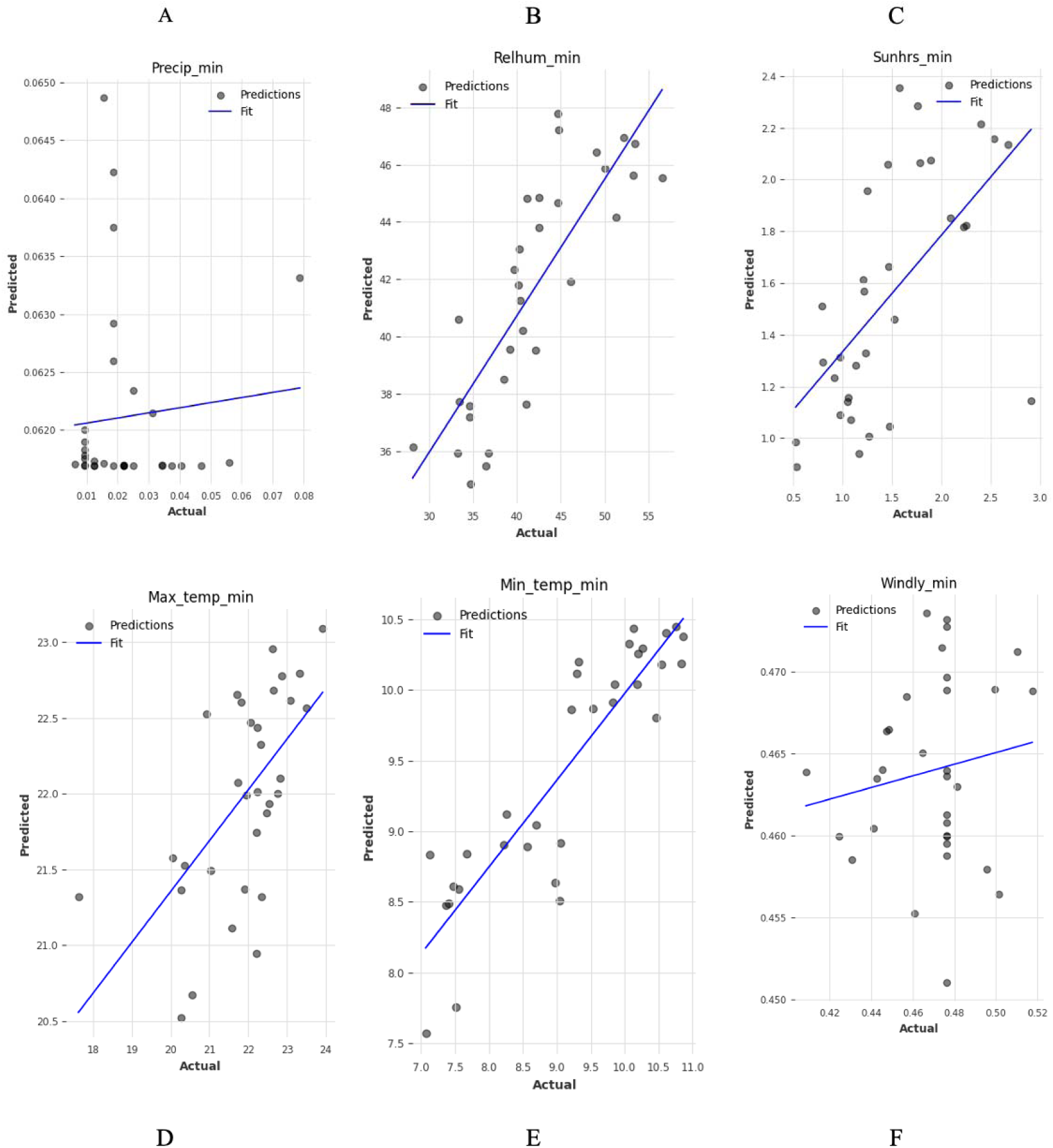
Actual vs. predicted values based on minimum trends. The black line indicates perfect prediction (y = x). Points clustered near the line indicate accurate predictions. A) LSTM achieved MAE=0.05& RMSE=0.05. B) Transformer achieved MAE=3.17 & RMSE=4.05. C) TCN achieved MAE=0.32 & RMSE=0.44. D) TCN achieve MAE=0.71 & RMSE=0.98. E) TCN scored MAE=0.47 & RMSE=0.56. F) TCN achieved MAE=0.02 & RMSE=0.02.

**Figure 5.**
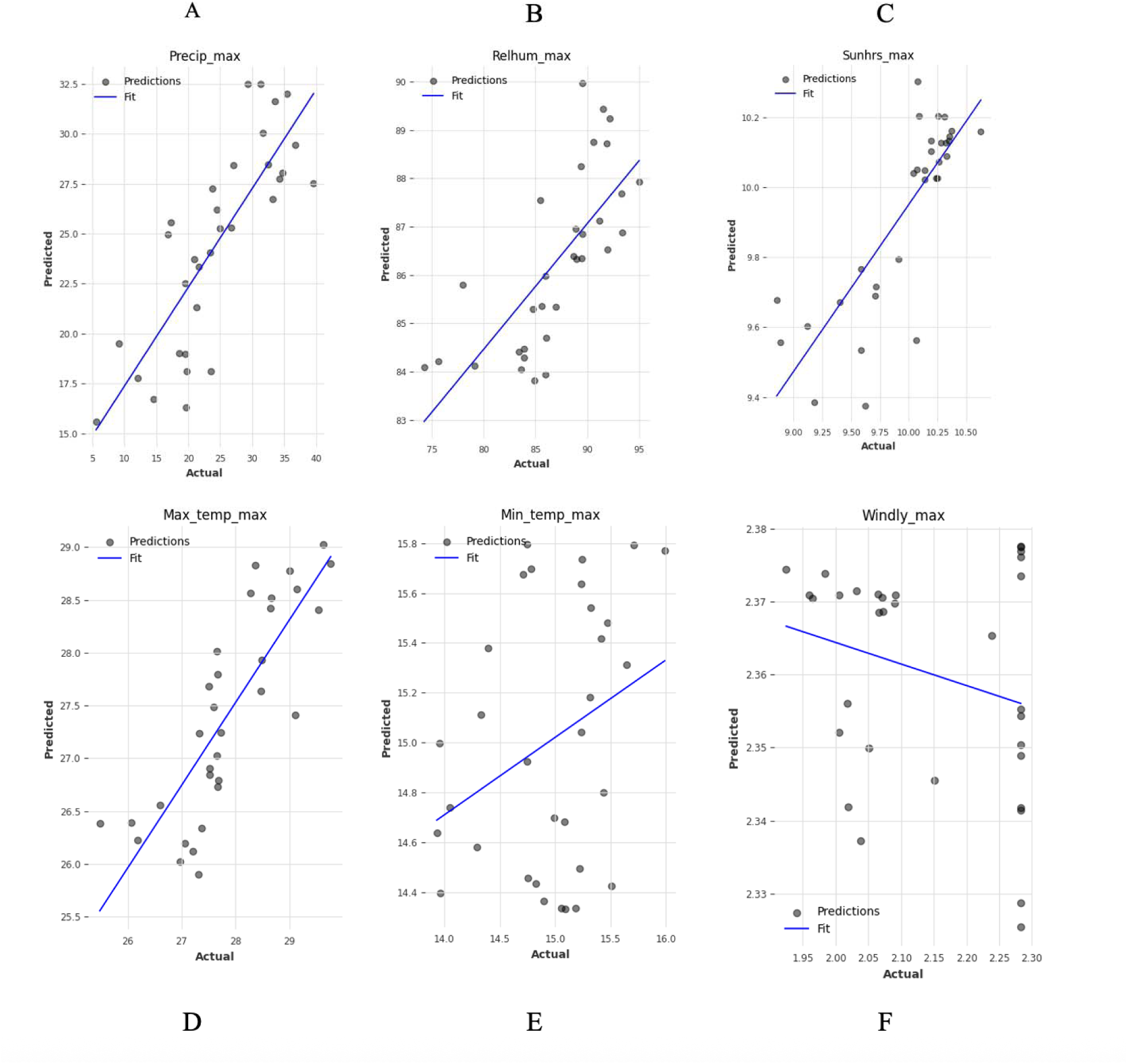
Actual vs. predicted values based on maximum trends. A) TCN scored MAE=4.29 & RMSE=5.11. B) TCN scored MAE=2.95 & RMSE=3.94. C) TCN achieved MAE=0.21 & RMSE=0.31. D) TCN score MAE=0.61& RMSE=0.74. E) TCN Scored MAE=0.39 & RMSE=0.47. F) Transformer achieved MAE=0.20 & RMSE=0.25. The black line indicates perfect prediction (y = x). Points clustered near the line indicate accurate predictions.

**Figure 6.**
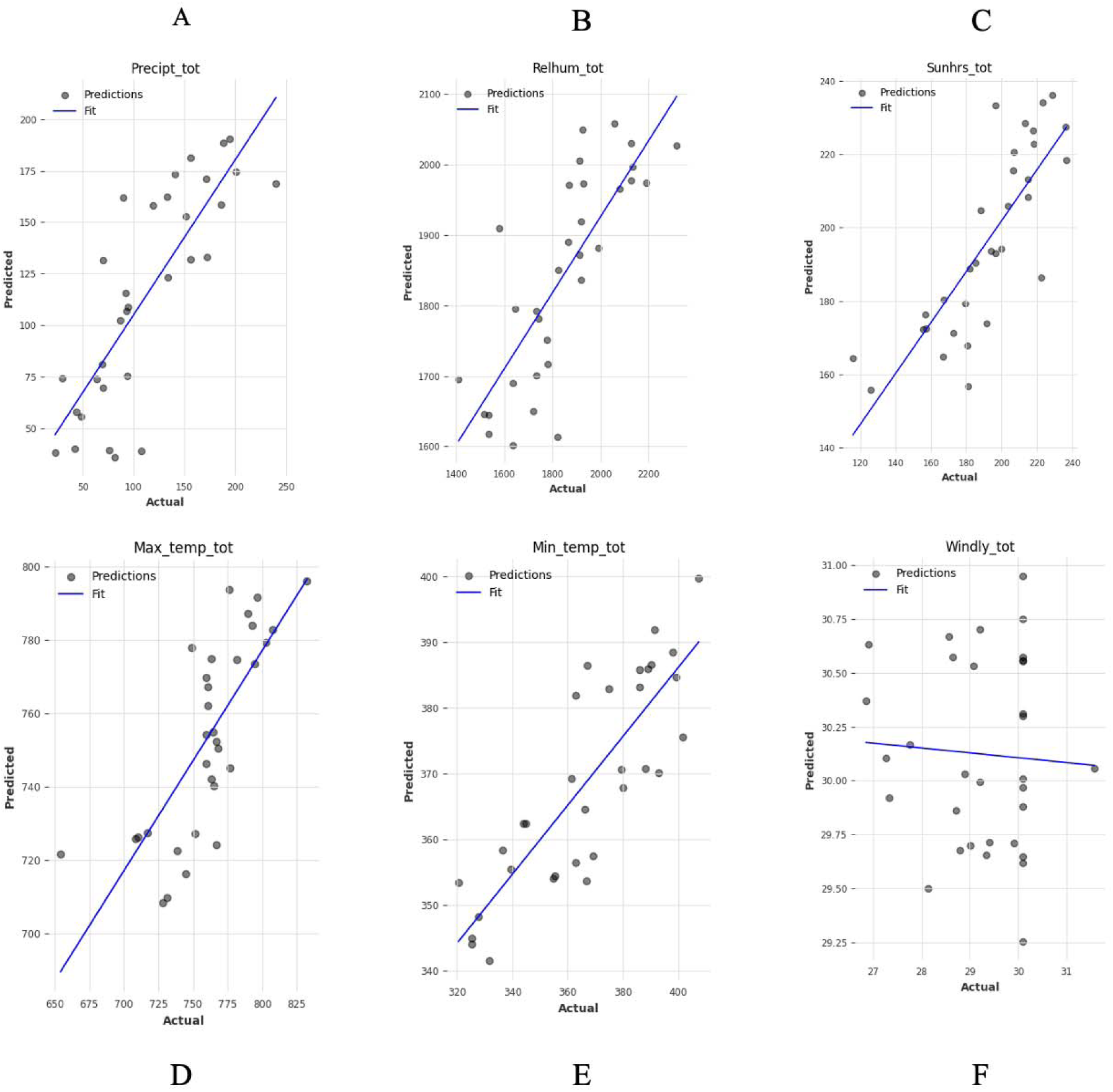
Actual vs. predicted based on total trends. A) LSTM achieved MAE=30.94 & RMSE=39.78. B) Transformer scored MAE=98.30 & RMSE=125.02. C) TCN scored MAE=12.77 & RMSE=16.73. D) TCN achieve MAE=18.84 & RMSE=22.87. E) TCN scored MAE=12.37 & RMSE=14.81. F) TCN achieved MAE=1.12 & RMSE=1.48. The black line indicates perfect prediction (y = x). Points clustered near the line indicate accurate predictions.

### Part II: Disease Prediction based on Climate Variables

#### Disease Incidence Prediction Performance

For both Malaria and Dysentery forecasting, the hurdle model performed better than the baseline in all dataset settings (minimum, average, maximum), and showed an improved R² (**Figure 7**). Both RMSE and MAE were lower with the hurdle model, showing improved accuracy and stability in continuous incidence predictions. Error bars demonstrated narrow variance across cross-validation folds, indicating robustness of the observed gains.

**Figure 7:**
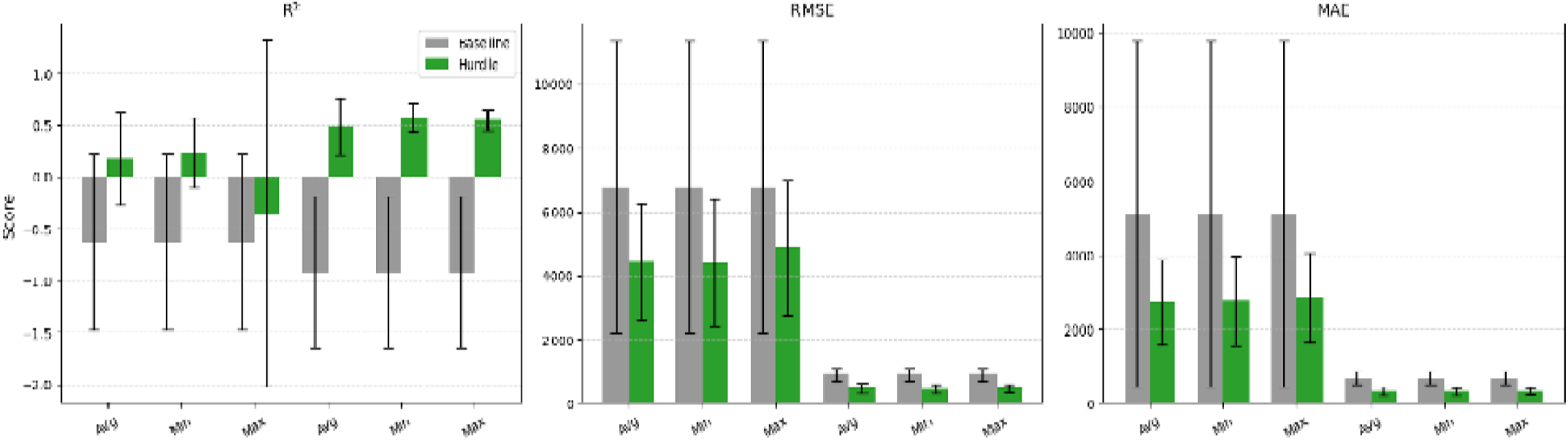
Incidence prediction performance across diseases (R², RMSE, MAE)

#### Disease Incidence Detection Accuracy

In incidence detection, the hurdle model achieved higher Precision, Recall, and F1-scores compared to the baseline across diseases and settings **(Figure 8).**

**Figure 8:**
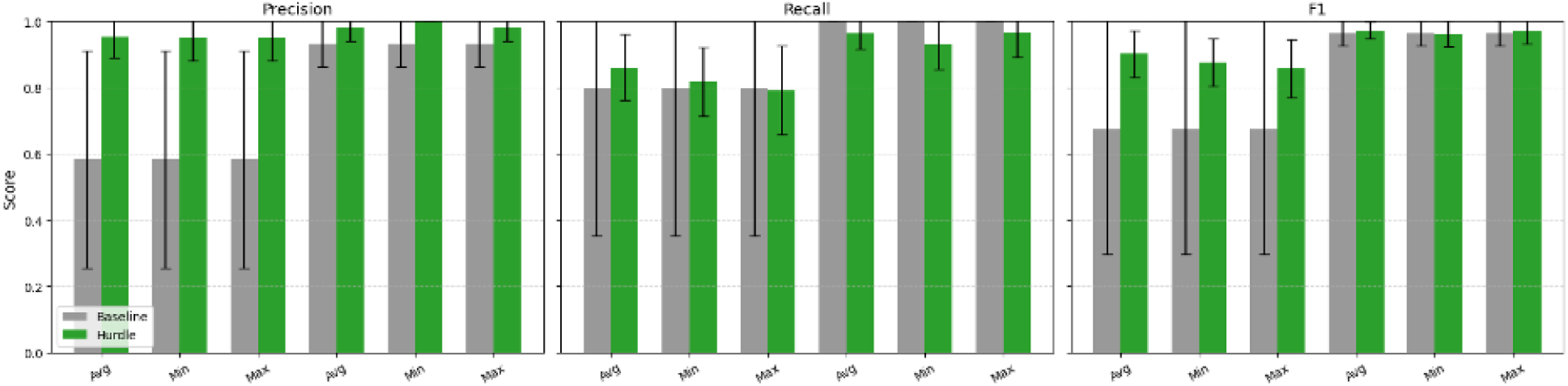
Overall Comparison of Incidence detection accuracy of diseases (Precision, Accuracy, F-Score)

#### Disease Incidence Error Analysis

Error stratification revealed that the hurdle model provided the greatest benefit during incidence periods, as indicated by substantially lower MAE for both incidence and non-incidence periods compared to th baseline (**Figure 9).**

**Figure 9:**
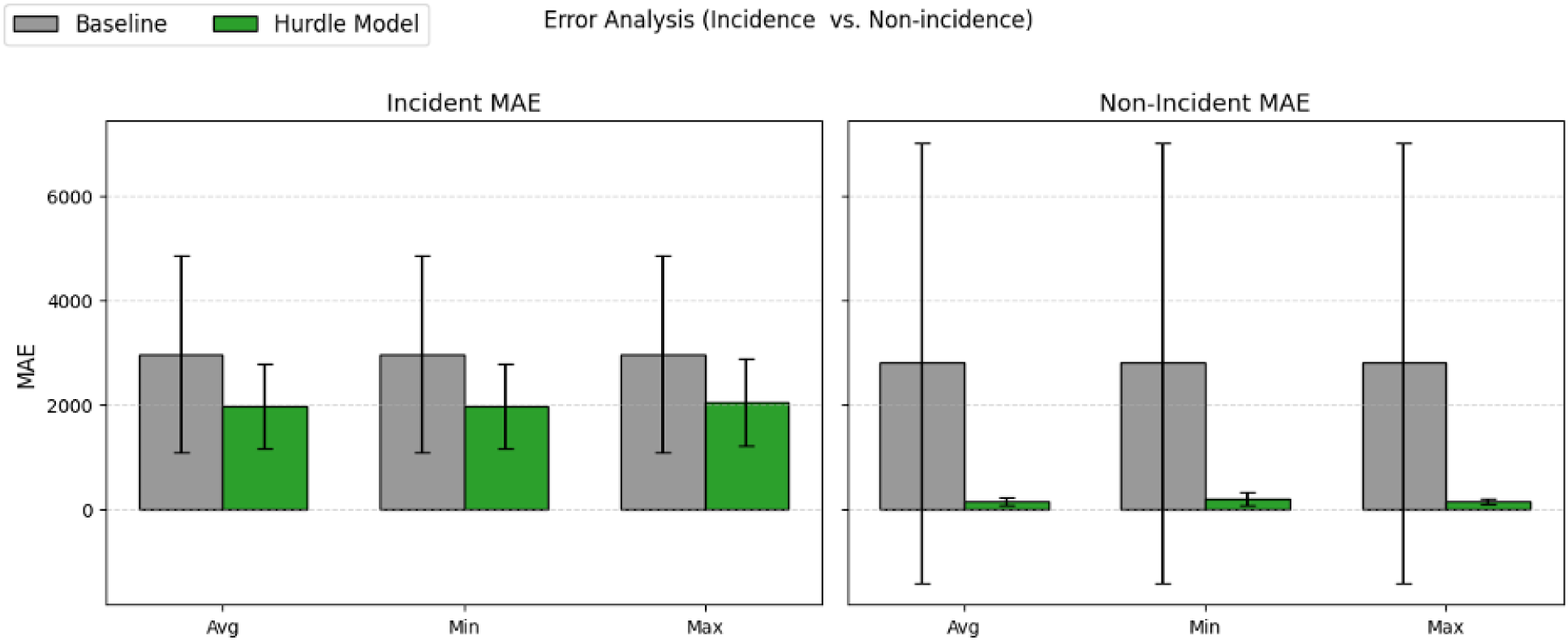
Error Analysis (Incidence vs. Non-Incidence)

### Model Selection and Proposed Climate-informed Disease Prediction Pipeline

As shown in Table 2, out of 72 evaluated experiments across four categories and six targets, the Transformer model achieved the highest number of statistically significant wins (n=18, 25.0%), followed by LSTM (n=9, 12.5%) and TCN (n=5, 6.9%) (Figure 10A). Category-level analysis (Figure 10B) indicates that the transformer model’s superiority is particularly pronounced in the categories’ minimum, maximum, and total. While TCN achieves the lowest average MAE, MSE, and RMSE across most of the variables and the best mean predictive performance, the Diebold–Mariano (DM) test reveals that the transformer model obtained the highest number of statistically significant wins according to the DM test across the six variables and 72 experiments. Therefore, based on aggregate performance and statistically significant improvements, our results support the selection of the transformer model as the primary architecture for disease incidence prediction (Figure 11). To illustrate the workflow of the best model, Figure 11 shows the model’s end-to-end processing pipeline: (I) the time-series input (Spatial-Climatic Features, covariates, and 24 Timesteps); (II) Spatial-Climatic Features time-series modeling using a transformer model performing inference; and (III) the disease incidence prediction pipeline (malaria and dysentery). In stage 1, incidence detection using XGBClassifier, and in Stage 2, XGBRegressor is used for incidence magnitude prediction, ultimately an input for decision support for real-time disease monitoring and actionable insight.

**Figure 10:**
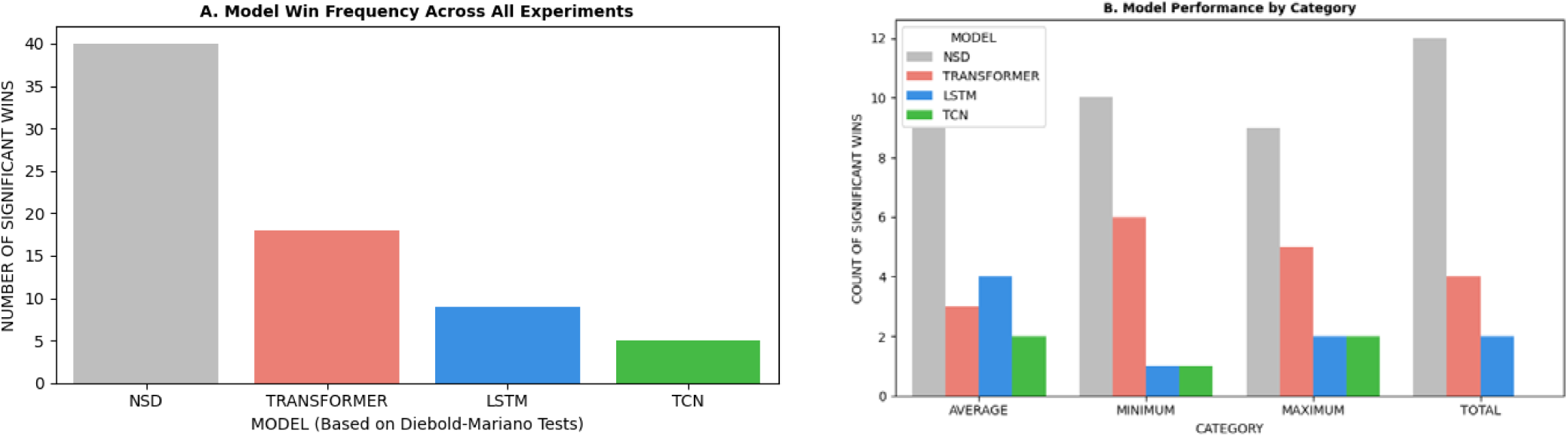
Aggregated Model Wins and Frequency Analysis A. Model Win Frequency Across All Experiments and B. Model Performance by Category

**Figure 11:**
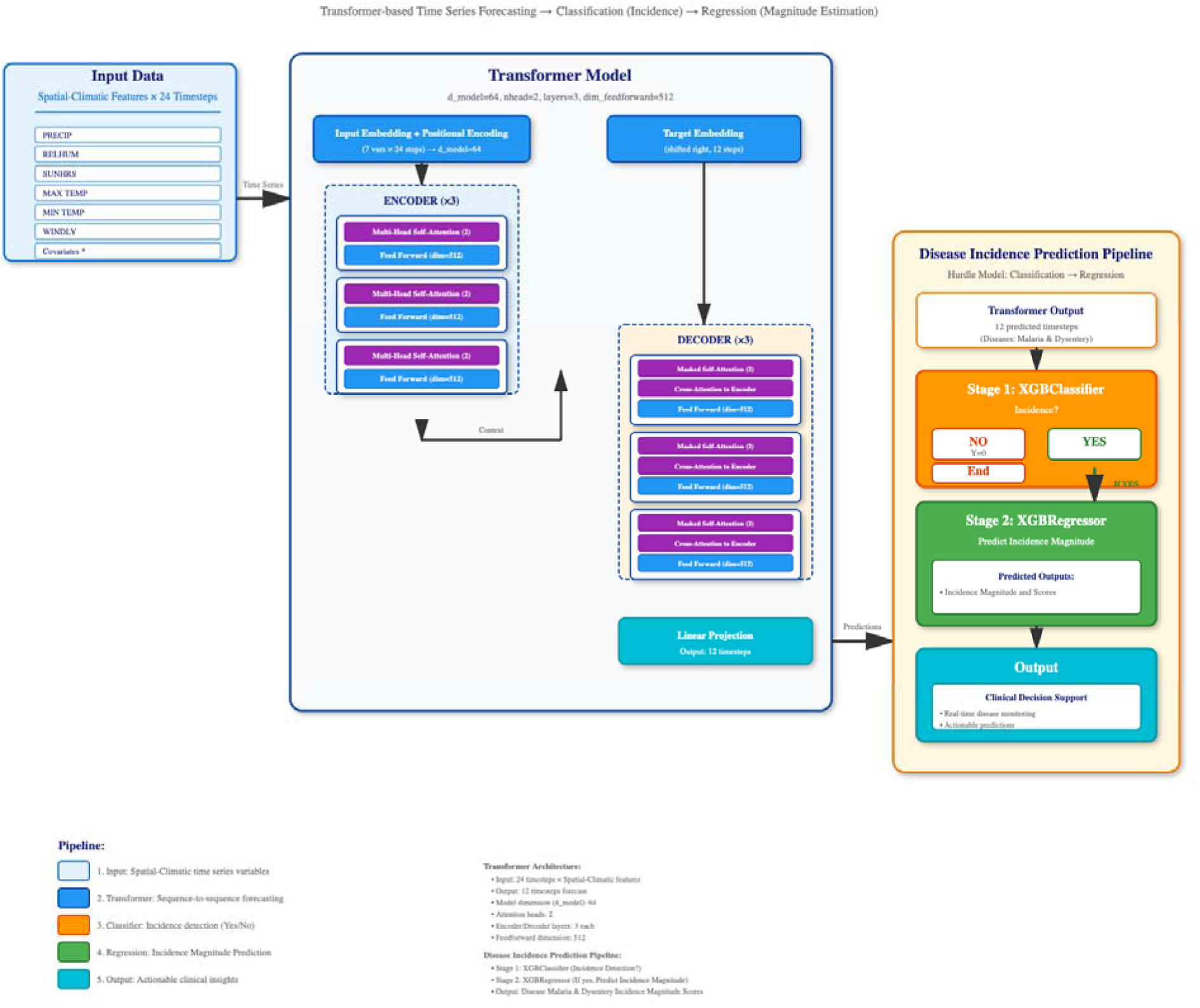
Climate-informed disease prediction pipeline: Time Series Forecasting, Classification (Incidence), and Regression (Magnitude Estimation)

## Discussions

In this study, we employed decadal climate and weather data, alongside spatial and temporal factors, to develop a climate-informed time-series deep learning model. The existing studies and models were: (I) based on abundant, standardized, statistically stable settings to predict future incidence using historical patterns and covariates, and (II) successful in learning continuous spatiotemporal representations rather than zero-inflated spatiotemporal representations. To address these gaps, the disease incidence problem was reformulated into a two-stage modeling pipeline to address extreme skewness (most months have zero or very low counts), rare but high-magnitude incidences, or a bimodal distribution (many zeros with occasional spikes). In the proposed two-stage forecasting architecture (Figure 2), the first model encodes weather variables, which a second-stage predictor then uses to estimate disease incidence. Overall, a modular climate-informed multi-disease prediction framework separates the task of climate feature model extraction from the tasks of disease incidence modeling to mitigate overfitting in low data settings, as well as to address the gaps between methodological proliferation marked by diverse architectures and epidemiological reality.

In recent years, the increasing availability of decadal climate and weather datasets has opened new opportunities for forecasting climate-sensitive infectious diseases. Studies have shown that deep learning architectures, including LSTM, Transformer, and CNN models, can outperform traditional statistical methods when meteorological variables (such as temperature, rainfall, and humidity) are included as inputs (47). At the same time, broader work on applying artificial intelligence (AI) to climate-health forecasting highlights the potential of machine learning (ML) to integrate environmental drivers but also notes key limitations in real-world epidemiological settings (48).

Most prior models fall into two broad categories: (I) models developed under relatively abundant, standardized, statistically stable conditions (for example full surveillance, high-quality data) that forecast future disease incidence based on historical patterns and covariates; and (II) architectures that focus on learning continuous spatio-temporal representations, rather than explicitly addressing the frequent zero-inflated, highly skewed, or “spike + zero” distributions observed in many endemic diseases (49). For example, many dengue-forecasting efforts assume a continuous incidence series rather than explicitly modelling the months of zero or near-zero cases (47).

However, in many low-resource and climate-sensitive settings, these assumptions often fail: surveillance data can be sparse or delayed, disease incidence may be extremely skewed (many months with zero or very low counts punctuated by rare but high-magnitude outbreaks), and end-to-end deep learning models trained directly on incidence may over-fit, generalize poorly, or fail to capture meaningful uncertainty (49).

To address this gap, we reformulate the disease-incidence forecasting problem into a two-stage modelling pipeline. First, a deep learning model is trained to forecast time-series weather and climate variables; second, those forecasts serve as inputs to downstream disease incidence prediction models (ranging from standard regression to hurdle frameworks designed to handle zero-inflation and spike dynamics). In the proposed two-stage forecasting architecture (Figure 2), the first stage encodes weather variables, and the second stage predicts disease incidence.

By separating climate feature extraction from disease incidence modelling, our modular “climate-informed multi-disease” prediction framework mitigates over-fitting in low-data settings, addresses the methodological versus epidemiological realism gap, and explicitly handles extreme outcome skewness and rare high-magnitude incidence periods. In our implementation, we systematically compare three deep-learning time-series architectures: LSTM, Transformer, and TCN, for weather forecasting across key environmental variables (precipitation, relative humidity, temperature, sunlight, wind speed), and then, in the second stage, apply hurdle modelling to forecast two diseases: malaria and dysentery. This two-stage design, where climate is forecast first and then disease incidence, departs from most existing studies, which directly forecast incidence from the history of incidence and covariates (47).

Ultimately, this work contributes to the climate-health modelling literature by offering a robust framework that (i) leverages decadal spatio-temporal climate data, (ii) handles zero-inflated, bimodal disease incidence distributions, (iii) disentangles climate prediction from disease prediction for modularity and generalization, and (iv) applies this design to multiple diseases in a low-resource context. The approach thus aims to deliver improved forecasting accuracy, actionable uncertainty quantification, and enhanced utility for public-health planning in climate-sensitive, data-sparse settings.

## Conclusion

This study presents a climate-informed, two-stage deep learning framework for spatio-temporal disease forecasting that integrates decadal climate and weather data with spatial and temporal features. The Transformer model achieved the highest number of statistically significant wins across all experiments, outperforming both LSTM and TCN in pairwise Diebold–Mariano tests. This suggests that the Transformer was the most robust model for capturing long-term dependencies in climate-variable forecasting. The second-stage hurdle model, designed for joint incidence classification and magnitude regression, achieved superior performance across all datasets for both malaria and dysentery. Error analysis confirmed that the hurdle framework reduced MAE during both incidence and non-incidence periods. Unlike conventional end-to-end models, the proposed modular pipeline first forecasts climate variables before predicting disease incidence, enhancing interpretability and generalization in data-sparse settings. Overall, this approach provides a scalable, climate-aware forecasting tool with potential applications in early-warning systems and public health planning, particularly in regions where climate-sensitive diseases are prevalent or evolving under changing environmental conditions.

## Ethical declarations

Approval for the research was granted by the ethical review board of the Jimma Institute of Technology, with the reference number RPD/JIT/172/15.

## Acknowledgements

The necessary resources for this study were provided by the AI and Biomedical Imaging Research Lab at Jimma Institute of Technology, and the Mega Research grant from Jimma University. We extend our gratitude to the Ethiopian Public Health Institute’s Public Health Emergency Management (PHEM) and the Ministry of Health of Ethiopia for granting us access to the valuable data required for this research.

## Ethics approval and consent to participate

This study was conducted in accordance with the principles of national and international guidelines for research involving human participants, as well as the principles outlined in the Declaration of Helsinki. Ethical approval for the research was granted by the ethical review board of the Jimma Institute of Technology, with the reference number RPD/JIT/172/15. The dataset is fully de-identified and contains only aggregated information. No individual-level identifiers are present, and no human subjects were directly involved in this research. All analyses were conducted in accordance with established ethical standards for secondary data research, including principles of privacy, confidentiality, and responsible data use.

## Consent for publication

Not applicable.

## Clinical trial number

Not applicable.

## Data availability

The datasets used and/or analyzed during the current study are available from the corresponding author upon reasonable request and with written permission from the original data sources.

